# The Acoustic Dissection of Cough: Diving into Machine Listening-based COVID-19 Analysis and Detection

**DOI:** 10.1101/2022.03.01.22271693

**Authors:** Zhao Ren, Yi Chang, Katrin D. Bartl-Pokorny, Florian B. Pokorny, Björn W. Schuller

## Abstract

**Purpose:** The coronavirus disease 2019 (COVID-19) has caused a crisis worldwide. Amounts of efforts have been made to prevent and control COVID-19’s transmission, from early screenings to vaccinations and treatments. Recently, due to the spring up of many automatic disease recognition applications based on machine listening techniques, it would be fast and cheap to detect COVID-19 from recordings of cough, a key symptom of COVID-19. To date, knowledge on the acoustic characteristics of COVID-19 cough sounds is limited, but would be essential for structuring effective and robust machine learning models. The present study aims to explore acoustic features for distinguishing COVID-19 positive individuals from COVID-19 negative ones based on their cough sounds.

**Methods:** With the theory of computational paralinguistics, we analyse the acoustic correlates of COVID-19 cough sounds based on the COMPARE feature set, i. e., a standardised set of 6,373 acoustic higher-level features. Furthermore, we train automatic COVID-19 detection models with machine learning methods and explore the latent features by evaluating the contribution of all features to the COVID-19 status predictions.

**Results:** The experimental results demonstrate that a set of acoustic parameters of cough sounds, e. g., statistical functionals of the root mean square energy and Mel-frequency cepstral coefficients, are relevant for the differentiation between COVID-19 positive and COVID-19 negative cough samples. Our automatic COVID-19 detection model performs significantly above chance level, i. e., at an unweighted average recall (UAR) of 0.632, on a data set consisting of 1,411 cough samples (COVID-19 positive/negative: 210/1,201).

**Conclusions:** Based on the acoustic correlates analysis on the COMPARE feature set and the feature analysis in the effective COVID-19 detection model, we find that the machine learning method to a certain extent relies on acoustic features showing higher effects in conventional group difference testing.

## Introduction

A novel coronavirus named severe acute respiratory syndrome coronavirus 2 (SARS-CoV-2) caused a disease that quickly spread worldwide at the end of 2019 and the beginning of 2020. In February 2020, the World Health Organization (WHO) named the disease COVID-19 and shortly after that declared the COVID-19 outbreak a global pandemic. Globally, as of February 2022, more than 434,150,000 confirmed cases of COVID-19, including more than 5,940,000 deaths were reported to the WHO^1^.

Both the presenting symptoms and the symptom severity vary considerably from patient to patient, ranging from asymptomatic infections or a mild flu-like clinical picture to severe illness or even death. Commonly reported symptoms of COVID-19 include (1) respiratory and ear-nose-throat symptoms such as cough, shortness of breath, sore throat and headache, (2) systemic symptoms such as fever, muscle pain, and weakness, as well as (3) loss of smell and/or taste (Esakandari et al., 2020). Less common ear-nose-throat symptoms associated with COVID-19 are pharyngeal erythema, nasal congestion, tonsil enlargement, rhinorrhea, and upper respiratory tract infection (El-Anwar et al., 2020).

### Diagnostic approaches

The early detection of a COVID-19 infection in a patient is essential to prevent the transmission of the virus to other hosts and provide the patient with appropriate and early treatment. A series of laboratory diagnosis instruments have been proposed to test for COVID-19, e. g., computed tomography (CT), real-time reverse transcription polymerase chain reaction (rRT-PCR) tests, and serological methods (Long et al., 2020; Tang et al., 2020). CT and X-ray detect COVID-19 based on chest images (Jiang et al., 2021; Raptis et al., 2020; Tabik et al., 2020; Zhang et al., 2021). An rRT-PCR test focuses on analysing the virus’ ribonucleic acid (RNA) and synthesised complementary deoxyribonucleic acid (cDNA) from a nasopharyngeal swab and/or an oropharyngeal swab (Rahbari et al., 2021). Serological instruments measure antibody responses to the corresponding infection and confirm the COVID-19 status (Tang et al., 2020). However, the instruments mentioned above are costly and/or not always available, since they can only be conducted by professional physicians and require special equipment. Even though rapid antigen and molecular tests are more and more used to quickly detect COVID-19, they often have instructions hard to follow for non-professionals, are not always accessible, and result in a huge amount of waste. It is essential to develop low-cost, real-time, easy-to-apply, and eco-friendly screening instruments that are ready-to-use everyday and basically everywhere.

### Disease detection based on bioacoustic signals

A promising approach for a screening tool fulfilling these requirements could be based on bioacoustic signals such as speech sounds or cough sounds (Brown et al., 2020; Hecker et al., 2021; B. W. Schuller, Batliner, et al., 2021; B. W. Schuller, Schuller, et al., 2021). Several studies have reported acoustic peculiarities in the speech of patients who have diseases associated with symptoms affecting anatomical correlates of speech production, such as bronchial asthma (Balamurali et al., 2020; Dogan et al., 2007) or vocal cord disorders (Falk et al., 2021; Jesus et al., 2015; Petrović-Lazić et al., 2011). Differences in various acoustic parameters were also found in recent studies comparing speech samples of COVID-19 positive and COVID-19 negative individuals (Asiaee et al., 2020; Bartl-Pokorny et al., 2021). Motivated by acoustic voice peculiarities found for various diseases, machine learning has been increasingly applied to automatically detect medical conditions from voice, such as upper respiratory tract infection (Albes et al., 2020), Parkinson’s disease (Yaman et al., 2020), and depression (Ringeval et al., 2019). Recent studies on the automatic detection of COVID-19 from speech signals achieved promising results through both traditional machine learning (Han et al., 2021; B. W. Schuller, Batliner, et al., 2021; Shimon et al., 2021; Stasak et al., 2021) and deep learning techniques (Hassan et al., 2020; Pinkas et al., 2020; B. W. Schuller, Batliner, et al., 2021). Although research on the automatic detection of diseases based on speech is rapidly expanding, it faces a number of challenges in terms of algorithm generalisability and potential application in real-world scenarios. These challenges include gender and age distribution, the presence of different mother tongues, dialects, sociolects, or cognitive aspects such as individual speech-language and reading competence that may affect various acoustic parameters (Alves et al., 2015; Goyal et al., 2021; Nagumo et al., 2020; Procter & Joshi, 2020; Rojas et al., 2020; Sun, 2020; Taylor et al., 2020). Studies on COVID-19 face additional challenges related to the fact that COVID-19 is a relatively new and not yet well understood disease with a wide range of symptoms and divergent symptom severity (Hu et al., 2020; Tu et al., 2020). Studies need to consider the symptom heterogeneity of COVID-19 positive patients and the fact that many symptoms may also occur in other diseases such as bronchial asthma or flu. Therefore, it is essential to consider the inclusion of patients with COVID-19-like symptoms but other diagnoses into COVID-19 negative study groups.

In contrast to speech, the acoustic parameters of cough sounds are less dependent on language-related aspects. Therefore, systems based on voluntarily produced cough sounds may be more easily applicable to a broader target group than speech-based systems. Cough is not only a promising bioacoustic signal since it reflects a body function performed by all people regardless of their culture or language competence, but is also one of the most prominent symptoms of COVID-19 and is closely related to the lung primarily affected by COVID-19.

### Physiology of cough

Cough is an important defence mechanism of the respiratory system as it cleans the airways through high-velocity airflow from accidentally inhaled foreign materials or materials produced internally in the course of infections. A cough is composed of an inspiratory, a compressive, and an expiratory phase (McCool, 2006). It is initiated with the inspiration of air (about 50% of vital capacity), followed by a prompt closure of the glottis and the contraction of abdominal muscles and other expiratory muscles. This process allows the compression of the thorax and the increase of subglottic pressure. The next phase of a cough constitutes the rapid opening of the glottis resulting in a high-velocity airflow (peak expiratory airflow phase), followed by a steady-state airflow (plateau phase) for a variable – voluntarily controllable – duration (Kelemen et al., 1987; Lee et al., 2021). The optional final phase is the interruption of the airflow due to the closure of the glottis (Lee et al., 2017). Cough can be classified into two broad categories: wet/productive cough with sputum excreted and dry/non-productive cough without sputum (Murata et al., 1998). Cough sounds were found to vary significantly due to a person’s body structure, sex, and the kind of sputum (Murata et al., 1998). Hashimoto and colleagues (Hashimoto et al., 2003) revealed that the ratio of the duration of the second phase to the total cough duration was significantly higher for wet coughs than for dry coughs. Chatrzarrin and colleagues (Chatrzarrin et al., 2011) compared acoustic characteristics of wet and dry coughs and found that the number of peaks of the energy envelope of the cough signal and the power ratio of two frequency bands of the second expiratory phase of the cough signal significantly differentiated between the two cough types.

### Disease detection from cough sounds

A number of researchers have been interested in potential acoustic differences between voluntarily produced cough sounds of patients with pulmonary diseases and healthy individuals. Knocikova and colleagues (Knocikova et al., 2008) compared the cough sounds of patients with chronic obstructive pulmonary disease (COPD), patients with bronchial asthma, and healthy controls. They found that patients with COPD had the longest cough duration and the highest power among the three groups. Higher frequencies were detected in the cough sounds of the bronchial asthma group compared with the COPD group. Furthermore, in the bronchial asthma group, the power of cough sound was shifted to a higher frequency range compared with the control group (Knocikova et al., 2008). Another study (Nemati, Rahman, Blackstock, et al., 2020) found that cough duration, MFCC1 (Mel-frequency cepstral coefficient), and MFCC9 features were the most important acoustic features for classification of pulmonary disease state (i. e., bronchial asthma, COPD, chronic cough, healthy) and disease severity, defined based on a patient’s forced expiratory volume in the first second (FEV1) divided through the forced vital capacity (FVC). Similar to the speech/voice domain, various automatic approaches have proved to be effective at detecting pulmonary diseases from cough sounds (Infante, Chamberlain, Kodgule, et al., 2017; Nemati, Rahman, Blackstock, et al., 2020); good performance was even achieved when differentiating between two obstructive pulmonary diseases, namely bronchial asthma and COPD (Infante, Chamberlain, Fletcher, et al., 2017). Furthermore, using acoustic features extracted from cough sounds, the study (Nemati, Rahman, Blackstock, et al., 2020) automatically classified the symptom severity of patients with pulmonary diseases. In another study (Sharan et al., 2018), cough sound analysis was used to predict spirometry results, i. e., FVC, FEV1, and FEV1/FVC, for patients with obstructive, restrictive, and combined obstructive-restrictive pulmonary diseases as well as healthy controls. Machine learning algorithms were also applied to distinguish pertussis coughs from croup and other coughs in children (Parker et al., 2014). The study (Nemati, Rahman, Nathan, et al., 2020) used a random forest algorithm to classify wet and dry coughs based on a comprehensive set of acoustic features and achieved an accuracy of 87%. Based on improved reverse MFCCs, the study (Zhu et al., 2016) achieved an accuracy in the classification of wet and dry coughs of 93.66% using hidden Markov models.

### COVID-19 detection based on cough sounds

A set of studies have investigated detecting COVID-19 from cough sounds. Alsabek and colleagues (Alsabek et al., 2020) compared MFCC acoustic features in cough, breathing, and voice samples of COVID-19 positive and COVID-19 negative individuals. They found a higher correlation between the COVID-19 positive group and the COVID-19 negative group for the voice samples than for the cough and breathing samples. Therefore, they concluded that the cough and breathing of a patient may be more suitable for detecting a COVID-19 infection than his or her voice. Another study (Cohen-McFarlane et al., 2020) collected cough sounds from public media interviews with COVID-19 positive patients and analysed them for the number of peaks present in the energy spectrum and power ratio between the first two phases of each cough event. They found the majority of cough sounds to have a low power ratio and a high number of peaks, a characteristic pattern previously reported for wet coughs (Chatrzarrin et al., 2011). Brown and colleagues (Brown et al., 2020) compared several hand-crafted features extracted from their collected crowd-sourced cough sounds of COVID-19 positive and COVID-19 negative individuals. They found that coughs from COVID-19 positive individuals are longer in total duration, and have more pitch onsets, higher periods, and lower root mean square (RMS) energy. In contrast, their MFCC features have fewer outliers compared to those of COVID-19 negative individuals.

The reported differences in acoustic features extracted from cough sounds of COVID-19 positive and COVID-19 negative individuals are promising for the automatic detection of COVID-19. To process hand-crafted features, traditional machine learning methods such as support vector machines (SVMs) and extreme gradient boosting were utilised (Brown et al., 2020; Mouawad et al., 2021; B. W. Schuller, Batliner, et al., 2021). End-to-end deep learning models were developed to detect COVID-19 from the log spectrograms of cough sounds, and performed better than the linear SVM baseline (Coppock et al., 2021). Similarly, deep learning was also successfully used to process MFCCs (Hassan et al., 2020; Laguarta et al., 2020; Pahar et al., 2021; Pahar & Niesler, 2021) or Mel spectrograms of cough sounds (Imran et al., 2020). The studies above have raised the potential and shown the effectiveness of machine learning for a cough sound-based detection of COVID-19.

### Contributions of this work

In the present study, we analyse acoustic differences in cough sounds produced by COVID-19 positive and COVID-19 negative individuals and further explore the feasibility of machine listening techniques to automatically detect COVID-19. On the one hand, we include COVID-19 positive and COVID-19 negative individuals irrespective of the presence or absence of any symptoms associated with COVID-19. On the other hand, this study aims to address the above-mentioned challenges of symptom heterogeneity of COVID-19 positive patients including non-symptomatic COVID-19 infections as well as similarities of symptoms to symptom characteristics of other diseases. Thus, we also investigate the isolated scenarios of COVID-19 positive and COVID-19 negative individuals all of which showing COVID-19-associated symptoms, and of COVID-19 positive and COVID-19 negative individuals all of which not showing any COVID-19-associated symptoms. Data for our experiments is taken from the open COUGHVID database (Orlandic et al., 2021a) that provides 27 550 cough recordings in conjunction with information about present symptoms. We analyse the acoustic features of the Computational Paralinguistics challengE (ComParE) feature set that recently achieved good performance for COVID-19 detection from cough sounds (B. W. Schuller, Batliner, et al., 2021). Furthermore, we train an effective machine learning classifier based on the extracted ComParE features. Finally, we investigate the contribution of the acoustic features extracted from the cough sounds to the COVID-19 status predictions of the machine learning classifier.

## Materials and Methods

### Databases and Tasks

#### Data Pre-processing

The dataset in our study is selected from the ongoing crowd-sourcing COUGHVID data collection (Orlandic et al., 2021a, 2021b). The COUGHVID database is collected via a web interface^2^. Thus, the recordings can be collected with personal computers, laptops, or smartphones. All cough recordings are voluntarily produced by the participants. The participants receive safe coughing instructions on the web page, e. g., holding the smartphone at arm’s length and coughing into the crook of the elbow, putting the phone into a plastic zip bag. Each audio recording lasts up to 10 seconds. At the time of analysis for this study, the latest released COUGHVID database consists of 27,550 cough sound files. There are three statuses of a cough sample for each participant to self-report: healthy, symptomatic without COVID-19 diagnosis, and COVID-19. Additionally, it is optional for each participant to report the geographic location (latitude, longitude), age, gender, and whether she/he has other pre-existing respiratory conditions and muscle pain/fever symptoms. Apart from the self-reported information, the data collectors trained an extreme gradient boosting (XGB) classifier on 121 cough sounds and 94 non-cough sounds to predict the probability of a recording containing cough sounds to exclude non-cough recordings (Orlandic et al., 2021a).

Since the participants with the symptomatic status did not explicitly report whether they were diagnosed with COVID-19, we only include the samples labelled as healthy (i. e., negative) and COVID-19 (i. e., positive) in the present study. Furthermore, we exclude samples with cough sound probabilities below or equal to 0.99 trying to ensure that each recording contains useful cough sounds. We note that no participant information was released in (Orlandic et al., 2021a). We assume the audio files with the same location, age, and gender were recorded from the same participant. To better implement subject-independent experiments, we guarantee that only one cough sound file per participant is included. Therefore, the audio files with the same location, age, and gender are reduced into a single one. Moreover, all audio files containing noise and speech are manually excluded. The final dataset for this study contains 1,411 audio files (3.75 h) with a mean duration of 9.57 ± 3.30 s standard deviation (SD); the audio files are re-sampled into 16 kHz.

For each status (i. e., COVID-19 negative, COVID-19 positive), four classes of clinical conditions are considered: no symptoms, respiratory symptoms only, muscle pain/fever symptoms only, and both aforementioned symptoms. For each symptom class, the total number of samples and their gender distributions are listed in Table 1. Similar to other COVID-19 related acoustic databases (Muguli et al., 2021; B. W. Schuller, Batliner, et al., 2021), the sample numbers at the two statuses are imbalanced. Furthermore, the age distribution is examined across all symptom conditions (see Figure 1).

**Table 1.** Total and gender-specific distribution of number of cough samples across COVID-19 status and symptom conditions. neg = COVID-19 negative, pos = COVID-19 positive, f = female, m = male, + = symptomatic, - = asymptomatic.

| Status |  | Symptoms | Total | Gender (f/m) |
| --- | --- | --- | --- | --- |
| neg | neg. | no | 996 | 293/703 |
|  | neg+ | respiratory only | 124 | 48/ 76 |
|  |  | muscle pain/fever only | 57 | 20/ 37 |
|  |  | both symptoms | 24 | 8/ 16 |
| Σ |  |  | 1 201 | 369/832 |
| pos | pos. | no | 111 | 36/ 75 |
|  | pos+ | respiratory only | 40 | 13/ 27 |
|  |  | muscle pain/fever only | 27 | 12/ 15 |
|  |  | both symptoms | 32 | 14/ 18 |
| Σ |  |  | 210 | 75/135 |

**Figure 1.**
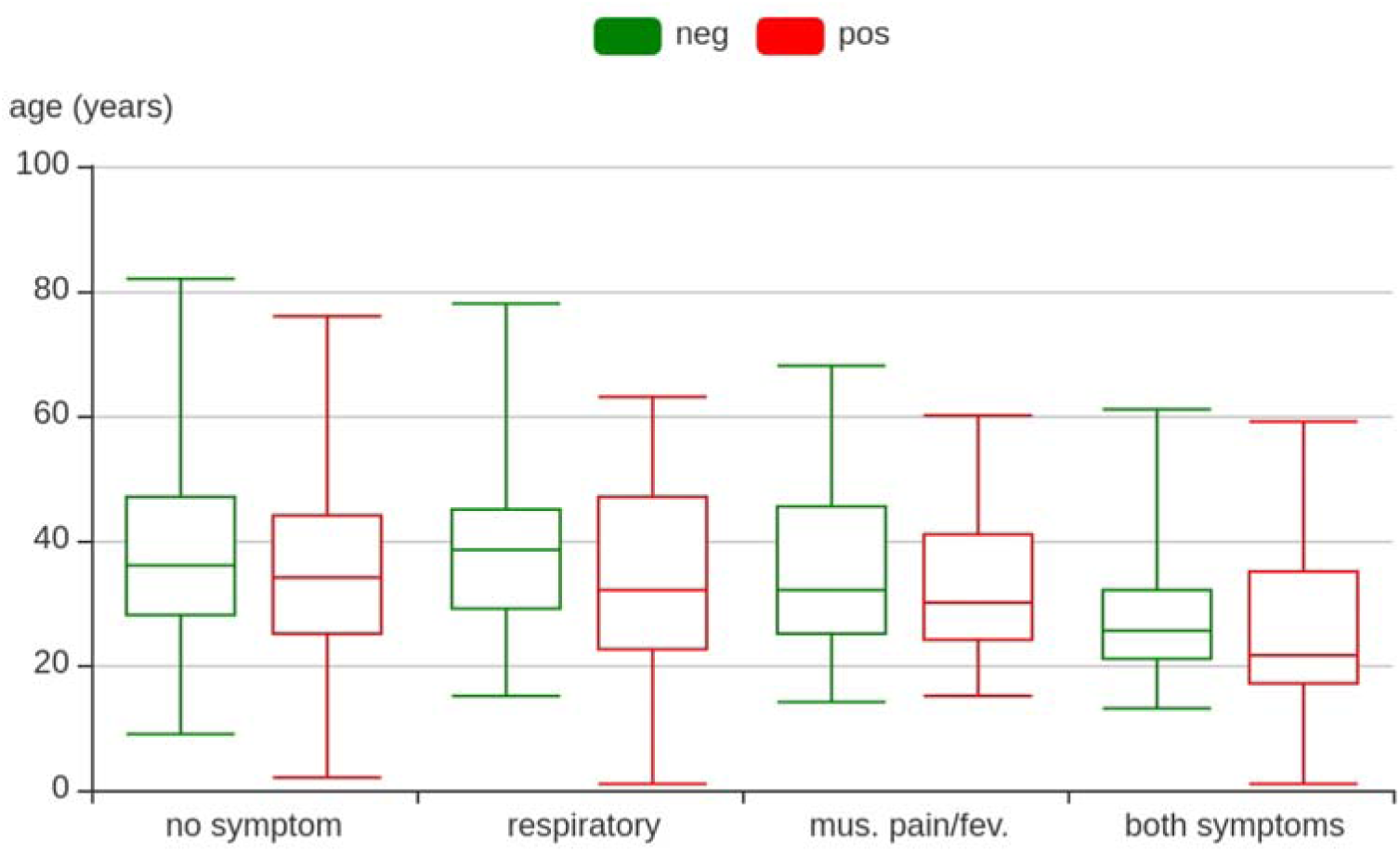
The age distribution of the 1,411 cough samples of the dataset for COVID-19 neg(ative) or (pos)itive. Mus.: muscle, fev.: fever.

#### Task Definition

This study has three major aims. Firstly, we compare acoustic features extracted from cough sounds of COVID-19 positive and COVID-19 negative individuals. Secondly, we apply machine listening methodology to detect COVID-19 based on cough samples. Thirdly, we focus on the explainability of automatic COVID-19 detection by comparing feature relevance in the machine listening approach to feature relevance according to the effect size in the group difference test. To achieve these aims, we group the dataset based on the related COVID-19 and symptom status into:

- samples of COVID-19 positive participants with respiratory and/or muscle pain/fever symptoms (pos_+_)
- samples of COVID-19 positive participants without respiratory and/or muscle pain/fever symptoms (pos_-_)
- samples of COVID-19 negative participants with respiratory and/or muscle pain/fever symptoms (neg_+_)
- samples of COVID-19 negative participants without respiratory and/or muscle pain/fever symptoms (neg_-_).

Based on these subgroups, we implement three clinically meaningful tasks:

- **Task 1: COVID-19 positive (pos) vs COVID-19 negative (neg)**. This task addresses the three aims of this study by including all samples of COVID-19 positive participants (210) and all samples of COVID-19 negative participants (1,201) irrespective of the presence or absence of symptoms.
- **Task 2: COVID-19 positive with symptoms (pos**_**+**_**) vs COVID-19 negative with symptoms (neg**_**+**_). This task addresses the three aims of this study by including only samples of COVID-19 positive participants with respiratory and/or muscle pain/fever symptoms (99) and samples of COVID-19 negative participants with respiratory and/or muscle pain/fever symptoms (205).
- **Task 3: COVID-19 positive without symptoms (pos**_**-**_**) vs COVID-19 negative without symptoms (neg**_**-**_). This task addresses the three aims of this study by including only samples of asymptomatic COVID-19 positive participants (111) and samples of asymptomatic COVID-19 negative participants (996).

Apart from all three tasks, it is interesting to investigate the performance in these tasks under different genders or age ranges, as previous studies have shown gender and age differences in cough behaviour (Bai et al., 2021; Ioan et al., 2014). As shown in Figure 1, the median of the age ranges is around 30, which is used to split the data into two age groups for each task.

### Acoustic Feature Extraction and Analysis

Both instrumental phonetic analysis and traditional machine learning build upon acoustic features in the cough signals. We extract features from every audio recording by the open-source toolkit openSMILE (Eyben et al., 2010) according to the ComParE feature set, which is the standard baseline feature set in the INTERSPEECH ComParE series (B. Schuller et al., 2014) and has proven effective in COVID-19 detection from cough sounds (B. W. Schuller, Batliner, et al., 2021). The ComParE feature set consists of 6,373 features calculated by several supra-segmental functionals, e. g., mean value, over segmental low-level descriptors (LLDs), e. g., loudness (→ mean (loudness) = mean of loudness). The LLDs, in the form of sequential features, are generated by analysing short-time segments, while the functionals focus on mapping the LLDs into a feature vector through computing statistical features inside each LLD and over multiple LLDs. The details of the ComParE feature set can be found in (B. Schuller et al., 2014).

Using the Kolmogorov-Smirnov test, we determine at a 5% significance level that most class-specific feature distributions are unlikely to come from standard normal distributions. Thus, we apply the non-parametric two-sided Mann-Whitney U test to analyse the extracted features for distribution differences between the COVID-19 positive and COVID-19 negative samples in each task. We further compute the effect size *r* as the correlation coefficient calculated as the *z*-value divided by the square root of the number of samples. Finally, we rank the features according to the effect size’s absolute value. Features showing at least a weak correlation effect (|r| ≥ .1) are considered relevant for the respective task. These features are referred to as top features.

### Automatic COVID-19 Detection

#### Classifiers

Based on all 6,373 extracted ComParE features, we apply machine learning methodology to study automatic COVID-19 detection in (three) binary classification tasks. Several classification approaches come into consideration, including linear models and non-linear models. A linear model learns a linear mapping between the inputs, i. e., the features, and the labels, i. e., the COVID-19 status; a non-linear model learns a non-linear mapping. In our work, a set of models are applied to detect COVID-19 from cough sounds. The employed linear models consist of linear regression models, i. e., least absolute shrinkage and selection operator (LASSO), Ridge, and ElasticNet, and a linear SVM model. Linear regression models, i. e., logistic regression in classification tasks, construct a linear model with different penalties, i. e., L1, L2, and a combination of L1 and L2, leading to the three models: LASSO, Ridge, and ElasticNet, respectively. The coefficients of the features in a linear regression model can be considered as the feature importance. Additionally, an SVM model is trained to find a hyperplane to maximise the margin between two classes. In linear SVM models, the coefficients of this hyperplane can be regarded as weights, whose absolute values indicate the relevance of each feature in the decision function – the larger the absolute value, the more important the respective feature (Chang & Lin, 2008; Guyon et al., 2002). Apart from the linear models, the utilised non-linear models contain decision tree, random forest, and multilayer perceptron (MLP). A decision tree constructs a tree-like model by learning simple decision rules from the features, and a random forest is composed of a number of decision trees for performance improvement. Both decision tree and random forest are able to calculate the feature importance, which is computed as the total reduction of the criterion brought by each feature. A set of hidden layers in MLP leads to a highly non-linear function between the inputs and the labels, which makes it challenging to interpret each feature’s role in the model. To learn the feature importance of neural networks (e. g., MLP), a set of methods have been proposed, such as deep learning important features (Shrikumar et al., 2017) and causal explanation (CXPlain) (Schwab & Karlen, 2019).

The ComParE features have shown effectiveness in various audio classification tasks, including pathological-speech-related disease detection (Cummins et al., 2020), on small to medium-sized datasets and represent one of the official machine learning pipelines of the INTERSPEECH ComParE series (B. Schuller et al., 2014; B. W. Schuller, Batliner, et al., 2021). In this study, we reapply this well-proven feature set in combination with the aforementioned models to investigate the basic feasibility of detecting COVID-19 from cough sounds. The whole ComParE features extracted from the audio samples are used as the input of the machine learning models.

Due to the limited size of the data, splitting the dataset into training, development, and test may lead to unreliable results. Hence, a five-fold cross validation strategy is used to generate the predictions over all audio samples. The whole dataset is equally split into five folds, each of which is used as the test set while the other four are employed as the training set. We combine all test sets in the 5-fold cross validation for performance evaluation; the final results are obtained on the combined dataset. The best hyperparameter is then selected corresponding to the best performance on the combined dataset. The linear regression models and the SVM model are optimised from multiple inverse values of regularization strength and complexity parameters C ∈ {10^−6^,10^−5^,10^−4^,10^−3^,10^−2^,10^−1^,1} respectively. Random forest is optimised from tree numbers in {50,100,150,200,250,300}. The decision tree is learnt with the default parameters in scikit-learn^3^. In the MLP model, three linear layers with the numbers of output neurons 1,024, 256, and 2. To avoid overfitting problems, the first two layers are followed by dropout operations with the probabilities of setting a neuron as 0.2 and 0.3, respectively. Notably, balanced class weights are applied to each SVM model in order to mitigate the data imbalance problem.

#### Evaluation Metrics

We use the unweighted average recall (UAR) to evaluate the classification performance purposefully without considering the data imbalance characteristics. UAR is the average of recalls on all classes. Additionally, we report the area under the receiver operating characteristic curve (AUC-ROC) calculated based on the probability estimates of each audio sample being predicted as the COVID-19 positive class. The AUC score may be inconsistent with the UAR, since the probability of a prediction is calibrated by Platt scaling and fit by an additional cross-validation procedure on the training data^4^. Finally, the confusion matrices for all three classification tasks are depicted to show the detailed performance.

#### Explainability of Automatic COVID-19 Detection

To provide an insight into the best performing linear classifier of each task, we export the respective model’s feature coefficients and calculate mean weights across all cross-validation folds. We then rank the features for relevance, i. e., according to the absolute value of the mean feature weights. The explainability of automatic COVID-19 detection via linear classification models is quantified by comparing feature relevance in the linear classification model to feature relevance according to the effect size in the non-parametric group difference test.

## Experimental Results and Discussions

### Results

#### Feature Analysis

The analysis of the extracted 6,373 ComParE features yields a number of features relevant for the investigated tasks. For Task 1, we identify 220 top features, i. e., features with an absolute value of the effect size *r* greater than or equal to .01 in the non-parametric group difference test. For Task 2 and Task 3, 1,567 and 46 features are found relevant, respectively. Table 2 reveals the LLDs underlying the respective top features of each task. All LLD categories of the ComParE set turn out to be relevant for the differentiation of symptomatic participants with and without COVID-19 (Task 2). However, when including asymptomatic participants (Task 1) or exclusively focusing on asymptomatic participants (Task 3), only selected energy-related, spectral, and voicing-related LLDs are found to differ at an absolute value of the effect size |r| ≥ .1. Figure 2 depicts the group-wise probability density estimates of the respective top one feature of each task, i. e., the mean inter-peak distance of the RMS energy with |r| ≥ .15 for Task 1, the mean inter-peak distance of the fourth MFCC with |r| ≥ .26 for Task 2, and again the mean inter-peak distance of the RMS energy with |r| ≥ .15 for Task 3. Fourteen out of 6,373 features are found to be jointly relevant for the differentiation between COVID-19 positive and COVID-19 negative in both symptomatic and asymptomatic participants (see Table 3).

**Table 2.** Categorisation of the 65 low-level descriptors (LLDs) of the COMPARE feature set and specification of involvement (√) or non-involvement (×) in a top feature of the respective differentiation task (Task 1: pos vs neg, Task 2: pos_+_ vs neg_+_, and Task 3: pos_-_ vs neg_-_). DDP = difference of differences of periods, HNR = harmonics-to-noise ratio, MFCC = Mel-frequency cepstral coefficient, neg = COVID-19 negative, pos = COVID-19 positive, RASTA = relative spectral transform, RMS = root mean square, + = symptomatic, - = asymptomatic.

| Group | Energy-related LLDs (4) | pos vs neg | pos<sub>+</sub> vs neg<sub>+</sub> | pos. vs neg. |
| --- | --- | --- | --- | --- |
| Prosodic | Auditory spectrum sum (loudness) | √ | √ | √ |
| Prosodic | RASTA-filtered auditory spectrum sum | √ | √ | × |
| Prosodic | RMS energy | √ | √ | √ |
| Prosodic | zero-crossing rate | × | √ | × |
| Group | Spectral LLDs (55) | pos vs neg | pos<sub>+</sub> vs neg<sub>+</sub> | pos. vs neg. |
| Cepstral | MFCC 1–14 | √ | √ | √ |
| Spectral | Psychoacoustic harmonicity | √ | √ | √ |
| Spectral | Psychoacoustic sharpness | × | √ | × |
| Spectral | Spectral centroid | × | √ | × |
| Spectral | Spectral energy 250–650 Hz, 1–4 kHz | √ | √ | √ |
| Spectral | Spectral entropy | × | √ | × |
| Spectral | Spectral flux | √ | √ | √ |
| Spectral | Spectral kurtosis | √ | √ | × |
| Spectral | Spectral roll-off point 0.25, 0.50, 0.75, 0.90 | √ | √ | × |
| Spectral | Spectral skewness | × | √ | × |
| Spectral | Spectral slope | √ | √ | √ |
| Spectral | Spectral variance | × | √ | × |
| Spectral | RASTA-filtered auditory spectral band 1–26 | √ | √ | √ |
| Group | Voicing-related LLDs (6) | pos vs neg | pos<sub>+</sub> vs neg<sub>+</sub> | pos. vs neg. |
| Prosodic | Fundamental frequency | × | √ | × |
| Quality | HNR | √ | √ | √ |
| Quality | Jitter (local and DDP) | × | √ | × |
| Quality | Shimmer | × | √ | × |
| Quality | Voicing probability | × | √ | × |

**Figure 2.**
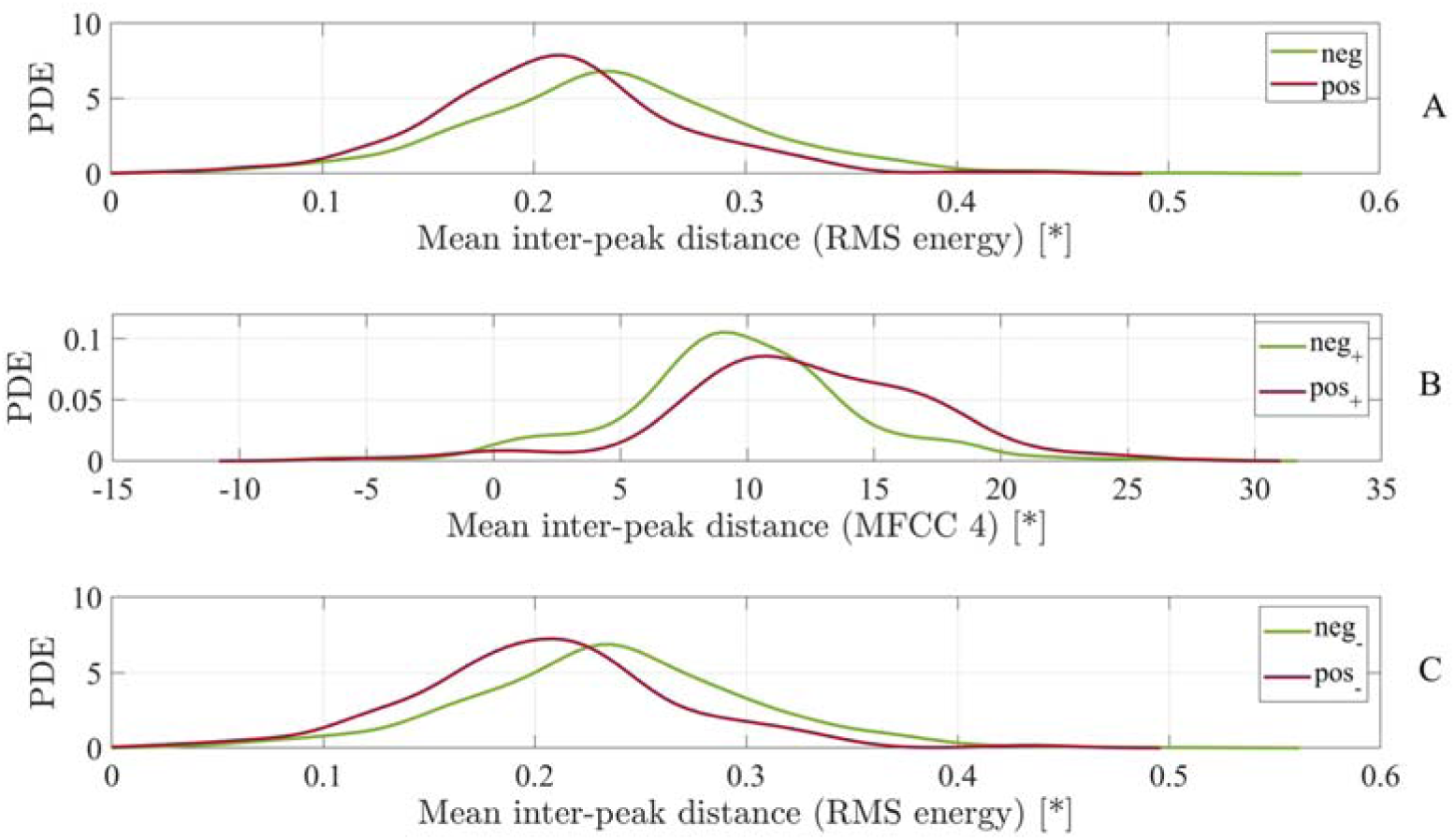
Comparison between part A: COVID-19 positive (pos) and COVID-19 negative (neg) participants (Task 1), part B: symptomatic COVID-19 positive (pos_+_) and symptomatic COVID-19 negative (neg_+_) participants (Task 2), and part C: asymptomatic COVID-19 positive (pos_-_) and asymptomatic COVID-19 negative (neg_-_) participants (Task 3) by means of the probability density estimate (PDE) of the top one feature of the respective differentiation task. MFCC = Mel-frequency cepstral coefficient, RMS = root mean square, * = real measurement unit does not exist as feature values refer to the amplitude of the digital audio signal.

**Table 3.** Joint top features between the pos_+_ vs neg_+_ (symptomatic COVID-19 positive vs symptomatic COVID-19 negative) and the pos_-_ vs neg_-_ (asymptomatic COVID-19 positive vs asymptomatic COVID-19 negative) differentiation tasks listed according to their mean ranks rounded to integers. HNR = harmonics-to-noise ratio, IQR = interquartile range, RASTA = relative spectral transform, RMS = root mean square, Δ= first-order derivative.

| Mean rank | Feature |
| --- | --- |
| 17 | Flatness ($\Delta$ spectral energy 250–650 Hz) |
| 19 | Flatness (spectral energy 250–650 Hz) |
| 27 | Flatness (RMS energy) |
| 33 | Flatness (spectralFlux) |
| 149 | Mean inter-peak distance (RMS energy) |
| 194 | Quartile 3 (HNR) |
| 195 | IQR 1–3 (HNR) |
| 196 | IQR 2–3 (HNR) |
| 202 | Mean inter-peak distance (loudness) |
| 291 | Mean inter-peak distance (spectral flux) |
| 410 | Skewness ($\Delta$ RASTA-filtered auditory spectral band 12) |
| 635 | Mean value of peaks (RMS energy) |
| 725 | Mean inter-peak distance (spectral harmonicity) |
| 786 | Mean value of peaks (loudness) |

#### Automatic COVID-19 Detection Results

The performance of the machine learning models for our three tasks is shown in Table 4. All of the best UARs on the three tasks achieve significant improvement over the chance (UAR: 0.5) (in a one-tailed z-test, pos vs neg: *p* < 0.001, pos_+_ vs neg_+_: *p* < 0.001, and pos_-_ vs neg_-_: *p* < 0.001. Correspondingly, the confusion matrices of all three results are depicted in Figure 3. All negative classes in the three tasks are classified with high true negative rates, while the true positive rates are around 0.5. Among the three tasks, for the task of pos_-_ vs neg_-_ we achieve the lowest UAR at the highest ratio of COVID-19 positive samples being incorrectly assigned to the negative class (see Figure 3). In Table 5, the performance on the data from the male participants is more superior than that from the female participants. Furthermore, the performance when ages are under or equal to 30 years is mostly better than those when ages are over 30 years.

**Table 4.** Classification performance in terms of unweighted average recall (UAR) and area under the receiver operating characteristic curve (AUC) for the three tasks. neg = COVID-19 negative, pos = COVID-19 positive, _+_ = symptomatic, _-_ = asymptomatic.

| Task |  | (1) pos vs neg |  | (2) pos+ vs neg+ |  | (3) pos- vs neg- |  |
| --- | --- | --- | --- | --- | --- | --- | --- |
| Samples (#) |  | 210/1,201 |  | 99/205 |  | 111/996 |  |
| Models |  | UAR | AUC | UAR | AUC | UAR | AUC |
| Linear | LASSO | 0.586 | 0.625 | 0.573 | 0.547 | 0.536 | 0.549 |
|  | Ridge | 0.632 | 0.671 | 0.653 | 0.641 | 0.558 | 0.594 |
|  | ElasticNet | 0.615 | 0.650 | 0.598 | 0.596 | 0.565 | 0.591 |
|  | SVM | 0.610 | 0.642 | 0.601 | 0.609 | 0.563 | 0.617 |
| Non-linear | Decision Tree | 0.521 | 0.521 | 0.538 | 0.538 | 0.501 | 0.501 |
|  | Random Forest | 0.500 | 0.651 | 0.544 | 0.606 | 0.500 | 0.600 |
|  | MLP | 0.558 | 0.600 | 0.593 | 0.606 | 0.505 | 0.570 |

**Figure 3.**
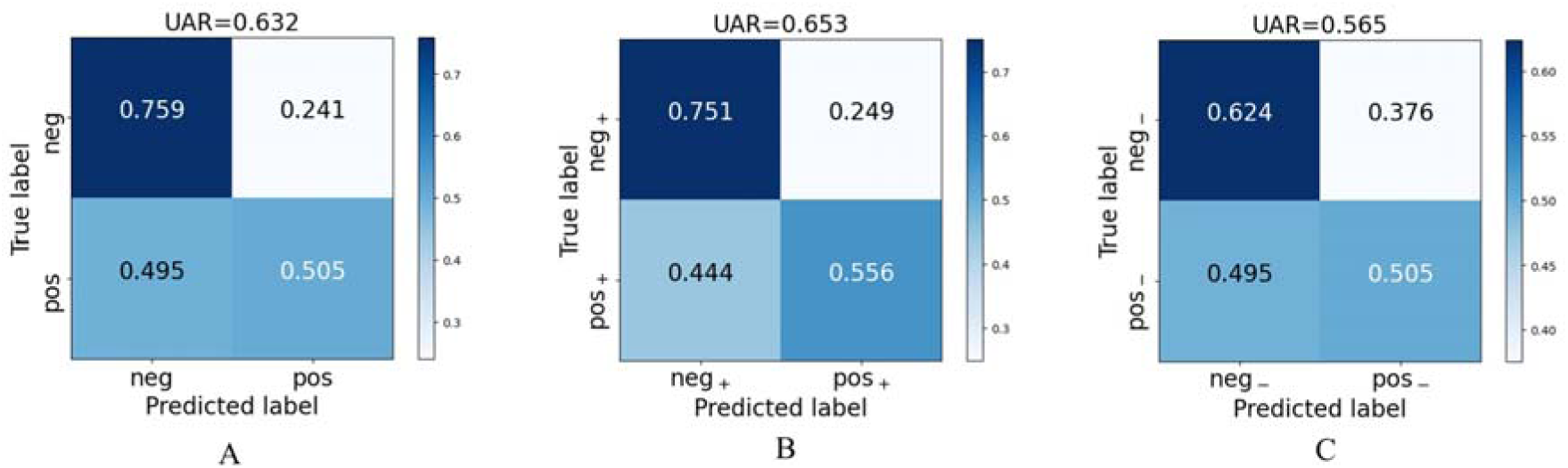
Confusion matrices for the three classification tasks complementary to the best results given in Table 4. Part A: (Task 1) pos vs neg, part B: (Task 2) pos_+_ vs neg_+_, part C: (Task 3) pos_-_ vs neg_-_. neg = COVID-19 negative, pos = COVID-19 positive, UAR = unweighted average recall, _+_ = symptomatic, _-_ = asymptomatic.

**Table 5.**
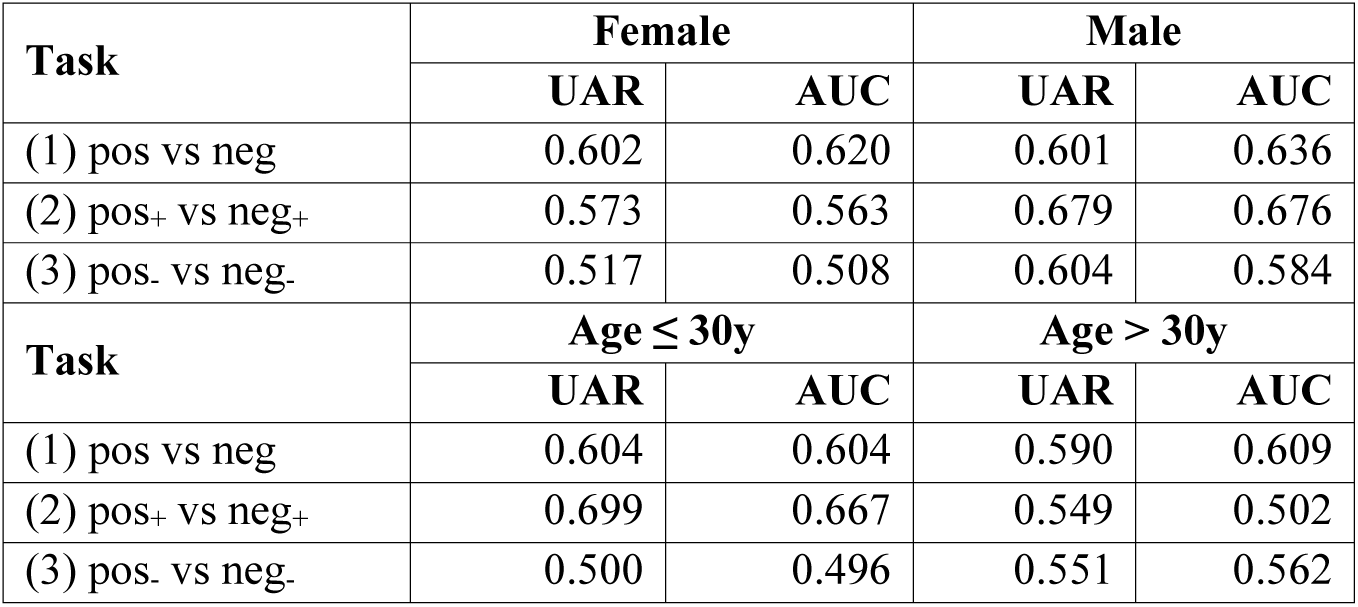
Gender-wise and age group-wise classification performance in terms of unweighted average recall (UAR) and area under the receiver operating characteristic curve (AUC) for the three tasks. The first two tasks (pos vs neg and pos_+_ vs neg_+_) are achieved by the Ridge models, and the third one (pos_-_ vs neg_-_) is achieved by the ElasticNet models. neg = COVID-19 negative, pos = COVID-19 positive, y = years, _+_ = symptomatic, _-_ = asymptomatic.

#### Explainability of the COVID-19 Detection Model

Our closer look at the weighting of acoustic features in the trained linear models reveals that identified top features according to the effect size in the non-parametric group difference test are also higher weighted in the respective best performing linear classification model for each task, i. e., Ridge for Tasks 1 and 2, and ElasticNet for Task 3 (see Figure 4). The 220 top features of Task 1 have a mean rank of 1,496 ± 1,640 SD amongst the features ranked according to the absolute value of the Ridge weights. Forty-nine out of the 220 top features are also among the first 220 Ridge weight ranked features. The 1,567 top features of Task 2 have a mean rank of 1,166 ± 1,084 SD amongst the Ridge weight ranked features. Herein, 1,173 out of the 1,567 top features are among the first 1,567 Ridge weight ranked features. The best performing model for Task 3, i. e., ElasticNet, only builds upon 250 non-zero feature coefficients. The 250 features with non-zero coefficients include 42 out of the 46 top features of Task 3. Eighteen out of the 46 top features are also among the first 46 ElasticNet weight ranked features.

**Figure 4.**
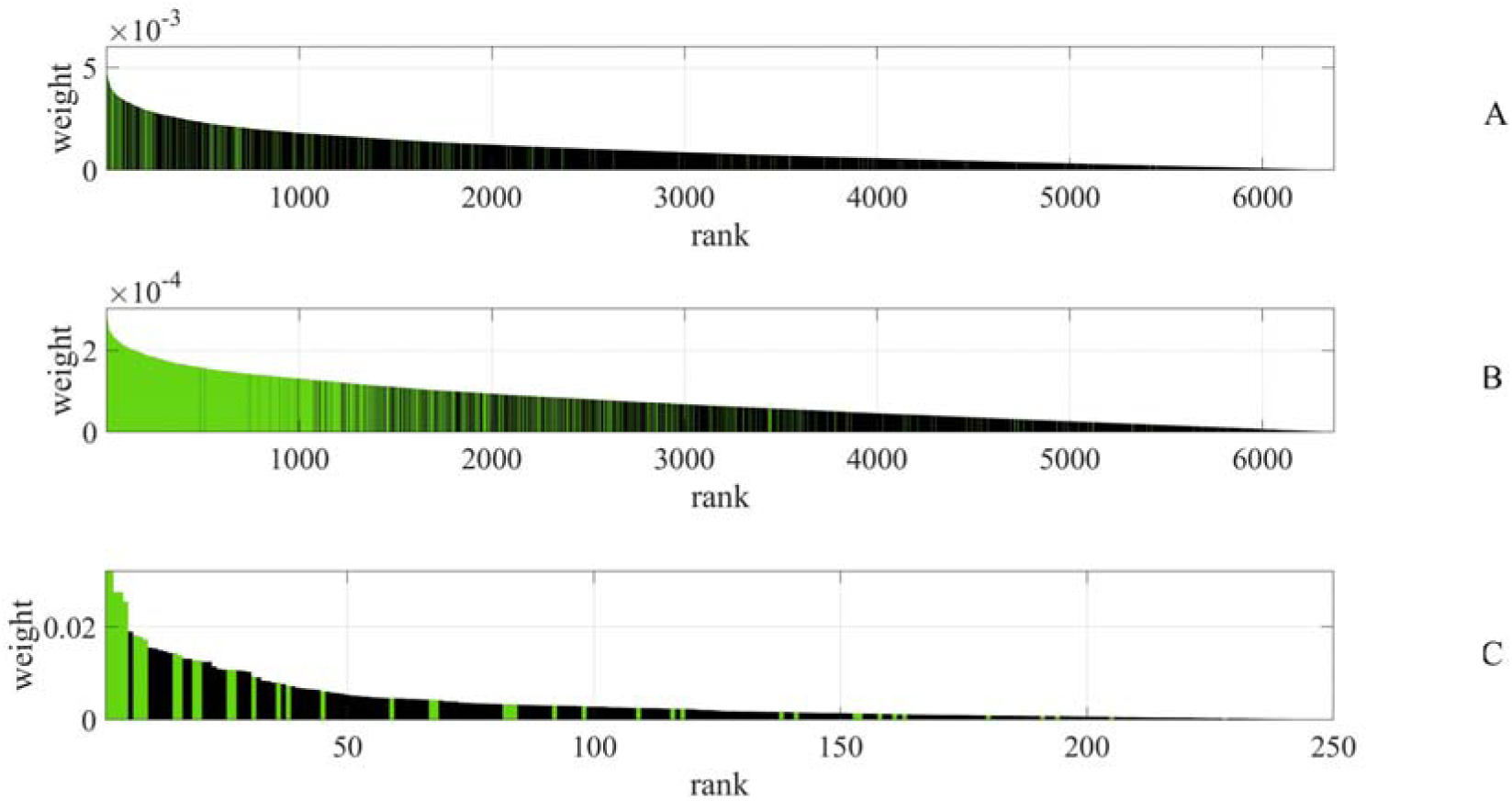
Feature ranking according to the absolute value of Ridge feature weights for part A: (Task 1) pos vs neg and part B: (Task 2) pos_+_ vs neg_+_, as well as of ElasticNet feature weights for part C: (Task 3) pos_-_ vs neg_-_. Green bars indicate top features according to the effect size in the non-parametric group difference test. A different x-axis scaling is used for (Task 3) as the ElasticNet model only builds upon 250 non-zero feature coefficients.

## Discussion

This study controls for the presence/absence of COVID-19-associated symptoms when comparing acoustic features extracted from cough sounds produced by COVID-19 positive and COVID-19 negative individuals and applying machine listening technology to detect COVID-19 automatically. Our study has important clinical implications as our results demonstrate the basic feasibility of automatic detection of COVID-19 based on cough sounds irrespective of the presence or absence of symptoms. Machine listening applications distinguishing between cough samples of symptomatic COVID-19 positive individuals and those of individuals with other diseases could advise the patient to stay at home and contact her/his doctor by phone before they go to the clinics/hospitals for medical professionals and may spread the virus there. By distinguishing between cough samples produced by asymptomatic COVID-19 positive and COVID-19 negative individuals, the easy-to-apply instrument can be developed to help prevent the unconscious transmission of the virus from asymptomatic COVID-19 positive individuals.

Although the classification performance of the SVM used in our study is significantly better than chance level, there are studies reporting better performances for COVID-19 detection based on cough sounds (Brown et al., 2020; Coppock et al., 2021). Brown and colleagues (Brown et al., 2020) studied three classification tasks: COVID-19/non-COVID, COVID-19 with cough/non-COVID with cough, and COVID-19 with cough/non-COVID asthma cough. The second and the third tasks were based on cough sounds and breath sounds, respectively, whereas the first task was based on both cough and breath sounds. All three tasks achieved over 80%, higher than those in our three tasks, which is possibly caused by two reasons. Firstly, the number of users in (Brown et al., 2020) is quite small and potentially less representative (Brain & Webb, 2000) as compared to our study: The number of users in (Brown et al., 2020) for each task is 62/220, 23/29, and 23/18, respectively, whereas the task-wise numbers of samples in our study are 210/1,201, 99/205, and 111/996. Secondly, the first and third tasks utilised breath sounds, which perhaps provide some discriminative features. Coppock and colleagues (Coppock et al., 2021) trained deep neural networks on the log spectrograms of both cough and breath sounds from the same crowd-sourced dataset as in (Brown et al., 2020) and achieved better results on the three tasks compared with the baseline, where SVMs processed the ComParE features. The AUCs in two of the three tasks are above 82% and the UARs are above 76%. Similarly, the better performance of this work could be caused by the limited number of participants (26/245, 23/19, and 62/293) and features from breathing sounds. In addition, an extra task of distinguishing COVID-19 and healthy participants without symptoms was set in (Coppock et al., 2021). Nevertheless, the performance on COVID-19 positive samples without symptoms was not reported independently. In contrast, such performance is evaluated in our Task 3, which is crucial for preventing COVID-19 transmission.

Our study reveals several acoustic peculiarities in COVID-19. As shown in Table 2, a set of LLDs could be helpful for differentiating COVID-19 positive individuals from COVID-19 negative ones. Across the three tasks, there are common LLDs of high relevance according to the effect size in the non-parametric group difference test, namely loudness, RMS energy, MFCCs, psychoacoustic harmonicity, spectral energy, spectral flux, spectral slope, RASTA-filtered auditory spectral bands, and HNR. Differences in RMS energy and MFCC-related features between the coughs of COVID-19 positive and COVID-19 negative individuals are also reported in (Brown et al., 2020). In our Task 2, all LLD categories of the ComParE set are found to bear relevant acoustic information to distinguish between the two groups. This might be due to an increased acoustic variability caused by symptomatic coughs, or due to the smaller number of samples available for this task. The analysed feature weights within the linear classification models show consistency with the features’ effect sizes, i. e., most top features according to the effect size also have higher weights in the linear classification models. This is a relevant finding towards the explainability of the applied machine learning approach. As indicated in Table 2, there are less top LLDs for Task 3. That might be because LASSO trends to use less features due to the nature of L1 regularisation. As a combination of LASSO and Ridge, ElasticNet is based on less features compared with Ridge.

As both speech and cough sounds are produced by the respiratory system, we herein compare and analyse peculiar acoustic parameters of patient’s speech and cough sounds. When analysing the acoustic peculiarities of patients with diseases that affect the anatomical correlates of speech production, the related studies reported that the peculiar acoustic parameters of the patients’ speech include fundamental frequency (*f*_o_), vowel formants, jitter, shimmer, HNR, and maximum phonation time (MPT) (Balamurali et al., 2020; Dogan et al., 2007; Jesus et al., 2015; Petrović-Lazić et al., 2011). Additionally, the peculiar acoustic parameters of voice samples of COVID-19 positive and COVID-19 negative participants were reported to include *f*_o_ standard deviation, jitter, shimmer, HNR, the difference between the first two harmonic amplitudes (H1–H2), MPT, cepstral peak prominence (Asiaee et al., 2020), mean voiced segment length, and the number of voiced segments per second (Bartl-Pokorny et al., 2021). We can find that there are common acoustic peculiarities between the voice of COVID-19 patients and patients with some other diseases: *f*_o_-related features, jitter, shimmer, HNR, and MPT. In Table 2, several acoustic LLDs of cough sounds have shown potential for distinguishing COVID-19 positive and COVID-19 negative individuals. Particularly, *f*_o_, jitter, shimmer, and HNR have high effective sizes in Task 2, i. e., pos_+_ vs neg_+_. The above findings indicate that there are similarities in acoustic peculiarities of speech and cough sounds of COVID-19 patients.

### Limitations

The classification performance reported in our study needs to be interpreted in the light of the well-known challenges of data collection via crowdsourcing, including data validity, data quality, and participant selection bias (Afshinnekoo et al., 2016; Khare et al., 2015; Porter et al., 2020). The COUGHVID database does not allow to verify the COVID-19 status of the participants, as the participants were not asked to provide a copy or confirmation of their positive or negative COVID-19 test. Another limitation is that the participants have not been instructed to record the data during a defined time window after the positive or negative COVID-19 test. Therefore, it is possible that some participants recorded their cough at the beginning of their infection, whereas others did the recording towards the end of their infection. Interestingly, the disease stage of COVID-19 was found to influence the nature of the cough (shifting from dry at an early disease stage to more wet at a later disease stage), concomitantly affecting acoustic parameters of the cough (Cohen-McFarlane et al., 2020). Moreover, the participants were asked to answer whether they had respiratory and/or muscle/pain symptoms, but no information on the severity of their symptoms is available. Although the safe recording instructions provided on the web page are reasonable with regard to the transmission of the virus, the suggestion to put the smartphone into a plastic zip bag while recording is suboptimal from an acoustic perspective. Another limitation of our study is that the participants did not receive clear instructions on how to cough, e. g., how often, or whether to take a breath between two coughs. Various audio recording devices and settings are inherent for crowdsourcing; we expect no bias towards one of the participant groups concerning the use of recording devices. We reduced files with the same location, age, and gender into a single one to ensure that only one cough sound file per participant is included, but we cannot guarantee that our dataset has only one sample per participant or that we have not mistakenly merged recordings from various individuals living in the same household.

Our target in this work was to explore the hand-crafted features’ importance for automatic COVID-19 detection. Some classifiers like k-nearest neighbours were not utilised as it might be difficult for them to output the feature coefficients/importance. Other approaches, such as transfer learning and end-to-end deep learning, were not used, as their inputs are either the original audio waves or simple time-frequency representations. Therefore, it is challenging to explain the features’ contribution with these methods. Additionally, we decided to apply a cross-validation schema due to the small dataset size, thus, testing was not entirely independent from the training as hyper-parameters were optimised on the test partitions.

Only the COUGHVID database was employed rather than multiple datasets in this study. Coswara (Sharma et al., 2020) was considered at the start of the experiments. However, the symptom information is not complete and well-organised for our study to analyse the effect of symptoms for detecting COVID-19. We also considered well-structured data, including University of Cambridge dataset collected by the COVID-19 Sounds app (Brown et al., 2020), diagnosis of COVID-19 using acoustics (DiCOVA) 2021 challenge data (Muguli et al., 2021), and INTERSPEECH ComParE 2021 challenge data (B. W. Schuller, Batliner, et al., 2021). However, these databases did not provide (sufficient) symptom information.

## Conclusions

In this study, we acoustically analysed cough sounds and applied machine listening methodology to automatically detect COVID-19 on a subset of the COUGHVID database (1,411 cough samples; COVID-19 positive/negative: 210/1,201). Firstly, the acoustic correlates of COVID-19 cough sounds were analysed by means of conventional statistical tools based on the ComParE set containing 6,373 acoustic higher-level features. Secondly, machine learning models were trained to automatically detect COVID-19 and evaluate the features’ contribution to the COVID-19 status predictions. A number of acoustic parameters of cough sounds, e. g., statistical functionals of the root mean square energy and Mel-frequency cepstral coefficients, were found to be relevant for distinguishing between COVID-19 positive and COVID-19 negative cough samples. Among several linear and non-linear automatic COVID-19 detection models investigated in this work, Ridge linear regression achieved a UAR of 0.632 for distinguishing between COVID-19 positive and COVID-19 negative individuals irrespective of the presence or absence of any symptoms and, thus, performed significantly better than chance level. Furthermore, the best performing machine learning models were found to rely to a certain extent on the acoustic features that yielded higher effects in conventional group difference testing.

## Outlook

From our point of view, it is highly important for future studies to specify the symptoms more clearly (e. g., severity estimates, onset time of symptoms), to include additional aspects potentially affecting the cough sound, such as smoking and vocal cord dysfunctions, and to differentiate in the COVID-19 negative group between participants with chronic respiratory diseases such as asthma or COPD and patients with a temporary infection such as the flu. Furthermore, it would be interesting for future studies to acoustically analyse the cough phases separately, as previous studies reported certain phase-specific acoustic peculiarities for wet and dry coughs (Chatrzarrin et al., 2011; Hashimoto et al., 2003). Moreover, it will be encouraging to consider more sound types (e. g., breathing and speech) and evaluate the physical and/or mental status of COVID-19 positive patients (e. g., anxiety) from speech for comprehensive COVID-19 detection and status monitoring applications (Han et al., 2020; Qian et al., 2021).

From the perspective of machine learning, feature selection methods will be investigated to extract useful features only. Deep learning models shall be explored for better performance due to their strong capability of extracting highly abstract representations. Particularly, when developing real-life applications for COVID-19 detection, it will be more efficient to skip the feature extraction procedure through training an end-to-end deep neural network with the input of audio signals or time-frequency representations. In addition to explaining linear classification models by analysing the weights of the acoustic features in this study, explaining deep neural networks along the dimension of time frame or frequency will need to be investigated to provide a detailed interpretation for each specific cough sound, i. e., when and at which frequency band a cough sound shows COVID-19-specific acoustic peculiarities. For this purpose, a set of approaches could be employed, e. g., local interpretable model-agnostic explanations (Ribeiro et al., 2016), shapley additive explanations (SHAP) (Lundberg & Lee, 2017), and attention mechanisms (Ren et al., 2020; Zhao et al., 2019).

## Supporting information

STROBE checklist

## Data Availability

The datasets (Orlandic et al., 2021a, 2021b) generated during and/or analyzed during the current study are available in the Zenodo repository, https://zenodo.org/record/4498364#.YekMM-rMLD4

## Acknowledgements

The authors want to express their gratitude to the holders of the COUGHVID crowdsourcing dataset for providing collected data for research purposes. Above all, thanks to all participants for their coughs. This work is supported by the European Union’s Horizon 2020 research and innovation programme under Marie Sklodowska-Curie Actions Initial Training Network European Training Network project TAPAS (grant number 766287), the Deutsche Forschungsgemeinschaft’s Reinhart Koselleck project AUDI0NOMOUS (grant number 442218748), and the Federal Ministry of Education and Research (BMBF), Germany, under the project LeibnizKILabor (grant No. 01DD20003).

## Footnotes

1 https://covid19.who.int/; retrieved 28 February 2022

2 https://coughvid.epfl.ch/; retrieved 8 October 2021

3 https://scikit-learn.org/stable/modules/generated/sklearn.tree.DecisionTreeClassifier.html

4 https://scikit-learn.org/stable/modules/svm.html#id11

